# A decade of cannabis-related hospital admissions in Victoria, Australia: A retrospective observational study

**DOI:** 10.64898/2026.07.25.26358914

**Authors:** Myfanwy Graham, Janneke Berecki-Gisolf, Jane Hayman, Adrian Carter, Shalini Arunogiri, Suzanne Nielsen

**Affiliations:** Monash Addiction Research Centre, Monash University, Melbourne, Australia; Victorian Injury Surveillance Unit, Monash University Accident Research Centre (MUARC), Monash University, Melbourne, Australia; School of Psychological Sciences, Monash University, Melbourne, Australia; Monash Bioethics Centre, School of Philosophy, History and Indigenous Studies, Monash University, Melbourne, Australia; Turning Point, Eastern Health, Melbourne, Australia

**Keywords:** Hospitalisations, cannabis, medical cannabis, delta-9-tetrahydrocannabinol, psychiatric disorders, psychosis

## Abstract

**Background and aims:** Over the past decade, there has been increased lifetime use of illicit and medical cannabis, underscoring the need to examine public health impacts. This included reports of increased cannabis-related psychosis. In this context, this study aimed to determine all cannabis-related hospital admissions over a ten-year period to characterise poisonings and mental health harms.

**Design:** This is a retrospective observational study of the Victorian Admitted Episodes Dataset (VAED) records, which include all hospital admissions in the state of Victoria and are supplied by the Victorian Department of Health.

**Setting:** Victoria, Australia.

**Cases:** Hospital records for poisoning (T40.7) and mental health and behavioural disorders (F12.0- F12.9) related to use of cannabis for the period July 2013 to June 2023 were selected for this study.

**Measurements:** Crude and age-standardised rates per 100,000 population are reported. Net percentage change over the ten-year study window was estimated using generalised linear models with a log-population offset, contrasting the final year (2022/23) against the baseline (2013/14).

**Findings:** There were 39,565 cannabis-related admissions in the ten-year period, including 7263 admissions where cannabis-related harm was the principal diagnosis. The age-standardised rate per 100,000 population was 63.6 for all cannabis-related harm admissions and 11.7 for principal diagnosis cases. The peak annual rate for all cannabis-related harm was observed in 2020/2021 (77.3 per 100,000). There was a 49.0% (p<0.0001) increase in the rate of hospital admissions with a cannabis-related principal diagnosis, when comparing 2022/23 with baseline (2013/14). Overall, harmful use (F12.1) was the most common cannabis-related mental and behavioural disorder. Psychotic disorder (F12.5) was the most common diagnosis (45.3%; n=2,909/6,421). When considering principal diagnosis only, the rates were highest in males aged 15-24 years (36.9/100,000), followed by females in the same age group (20.5/100,000). While most principal diagnosis cases involved males, a greater percentage change between the first and last years of the study period was observed for females (+75.1%, p<0.0001) compared with males (+34.6%, p=0.003).

**Conclusions:** A 49% increase in the rate of cannabis-related hospital admissions, particularly mental and behavioural disorders due to use of cannabinoids, has occurred over the past decade. These trends are non-linear, rates in the last two years were lower than the peak in 2020/2021.

**Declarations of competing interest:** Myfanwy Graham is an appointed member of the Therapeutic Goods Administration’s Medicinal Cannabis Expert Working Group. This article does not represent the views of the TGA or the Expert Working Group.

**Primary funding:** The Victorian Injury Surveillance Unit is supported by the Victorian Government. Myfanwy Graham is supported by an NHMRC Postgraduate Scholarship (GNT2030765) and a Monash Research Excellence Scholarship. Adrian Carter is supported by an Australian Research Council Future Fellowship (FT220100509). Suzanne Nielsen is an NHMRC Leadership Fellow (GNT2025894).

## INTRODUCTION

At a federal level in Australia, medicinal cannabis legalisation occurred in 2016, with rapid increases in medicinal cannabis prescription approvals over time (1). Concurrently, illicit cannabis remains the most commonly used illicit drug. Over the past decade, lifetime cannabis use and social acceptability have increased (2, 3). Use trends vary by population group, with notable increases in past year and high-risk use by females (4) (See Supplementary Figures S1 and S2).

Media reports have indicated that clinicians are concerned about increased cannabis-related hospital presentations and admissions related to prescribed medicinal cannabis (5), although there are limited peer-reviewed literature sources that have formally analysed these trends. There have also been clinician reports of cases of cannabis-related harm in Australia, including psychosis and cannabis hyperemesis syndrome (6–9). These signals of harm underscore the need to better understand the outcomes of increased population use, including unfavourable effects.

Hospital admission data provide insights into the impact of cannabis-related adverse events at a population level, alongside quantifying the health system burden. Inpatient hospitalisations provide insights into cases that may be more severe than acute ambulance attendances and emergency department presentations and therefore are ideal for examining the most severe consequences related to cannabis use (10). Examining trends in cannabis-related hospital admissions can also facilitate the identification of populations who may benefit from public health interventions aimed at reducing harm.

Considering these signals of increased cannabis use and related harm, this study aimed to quantify cannabis-related poisoning and mental health and behavioural disorder hospitalisations. Specifically, we aimed to determine crude and age-adjusted rates and trends in cannabis-related hospitalisations over a ten-year period.

## METHOD

This is a retrospective observational study of the Victorian Admitted Episodes Dataset (VAED).

### Design and Setting

VAED data were sourced by the Victorian Injury Surveillance Unit from the Victorian Department of Health. The VAED contains episode-level records for all public and private hospital admissions in Victoria, Australia, including demographic information, admission details, and diagnoses coded using the *International Statistical Classification of Diseases and Related Health Problems, 10th Revision, Australian Modification (ICD-10-AM*). This is a representative, near-complete dataset that reflects all admitted episodes for the period. For the calculation of population based rates, Victorian residential population estimates for each study year, by age group and sex, were sourced from the Australian Bureau of Statistics.

### Cases

The sample included hospital admission records for the period 1 July 2013 to 30 June 2023 that contained any cannabis-related ICD-10-AM diagnosis codes (T40.7; F12.0–F12.9). Two case groups were defined:

1. Poisoning-related cannabis cases, identified using the external cause code T40.7 (poisoning by narcotics and psychodysleptics [hallucinogens] - cannabis and cannabinoids).
2. Mental and behavioural disorders due to cannabinoid use, identified using diagnostic codes F12.0–F12.9.

To prevent double counting of hospital episodes, statistical transfers (changes of care type within the hospital) and other transfers were excluded. As required to preserve data confidentiality, cases with sex other than male or female were excluded, as these were small in number (average fewer than five per year over the study period).

Cases in which the cannabis-related ICD-10-AM code was the principal diagnosis (i.e. first-listed in the admission record) were selected and labelled as such. Mental and behavioural disorders due to cannabinoid use was further categorised as: acute intoxication (F12.0); harmful use (F12.1); dependence syndrome (F12.2); withdrawal state (F12.3); withdrawal state with delirium (F12.4); psychotic disorder (F12.5); amnestic syndrome (F12.6); residual & late onset psychotic disorder (F12.7); other mental and behaviours disorders (F12.8); unspecified mental and behavioural disorder (F12.9).

Age was provided in five-year age bands and further aggregated into child (0-14 years), adolescent (15-24 years), adult (25-64 years) and older adult (65+ years) groups.

### Measurements

Descriptive analyses were conducted to summarise cannabis-related admissions across the study period. For each outcome, annual age-standardised rates per 100,000 population were calculated using the direct method (11), with the Victorian standard population (2001) as the reference population. Age-standardisation was also applied within each broad age category (child, adolescent, adults, older adults).

### Statistical Analysis

To examine changes over time in cannabis-related hospital admission rates, Poisson regression models with a log link function were used. Annual counts of cannabis-related hospital admissions were modelled as a function of financial year, age group, and sex. Because the outcome was a rate, the log of the population at risk for each stratum (age group × sex × year) was included as an offset term to estimate rate ratios. To account for overdispersion, a quasi-Poisson model was used. The scale parameter was estimated empirically from the data using the deviance divided by the degrees of freedom (via the dscale option in SAS PROC GENMOD), rather than being fixed at 1. The financial year was modelled as a categorical variable, and the change in rate over the study period was estimated by contrasting the last year of data (2022/23) with the first year of data (2013/14), which was set as the reference period. An estimate statement was used to obtain the exponentiated coefficient for 2022/23 vs. 2013/14, which was then converted to a percentage change in admission rates. Analyses were conducted using SAS 9.4.

### Ethics

The Victorian Injury Surveillance Unit has Monash University Human Research Ethics Committee approval for the custody, use and dissemination of Victorian hospital admissions and Emergency Department data, project ID 44606.

## FINDINGS

### Cannabis-related hospital admissions

Between 1 July 2013 and 30 June 2023, there were 39,565 hospital admissions with a cannabis-related diagnosis. Overall, the average annual age-standardised rate per 100,000 population was 63.6 for all cannabis-related harm admissions and 11.7 for principal diagnosis cases (see Appendix A1). Over the ten-year study period, there was a significant net increase in the rate of cannabis related hospital admissions. Compared with the baseline year, the adjusted rate of admissions in 2022/23 was 50.4% higher than in 2013/14 (95% CI: 35.6%, 66.7%; p < 0.001). Similarly, for principal diagnosis cases only, the rate in 2022/23 was 49.0% higher than in 2013/14 (95% CI: 26.5%, 75.6%; p < 0.001).

### Cannabis-related hospital admission age-standardised rate by diagnosis

The 39,565 cannabis-related hospital admissions included n=37,821 cases with a diagnosis in the range F12.0-F12.9 and n=1,859 with a cannabis poisoning diagnosis (T40.7). One hundred and fifteen cases had both a poisoning and an F code diagnosis. Cannabis-related harm was the principal diagnosis in 7,263 (18.4%) admissions. During the ten-year period, cannabis-related hospital admissions peaked at two time points: 2017/18 (72.4 per 100,000) and 2020/21 (77.3 per 100,000) (see Figure 1).

**Figure 1.**
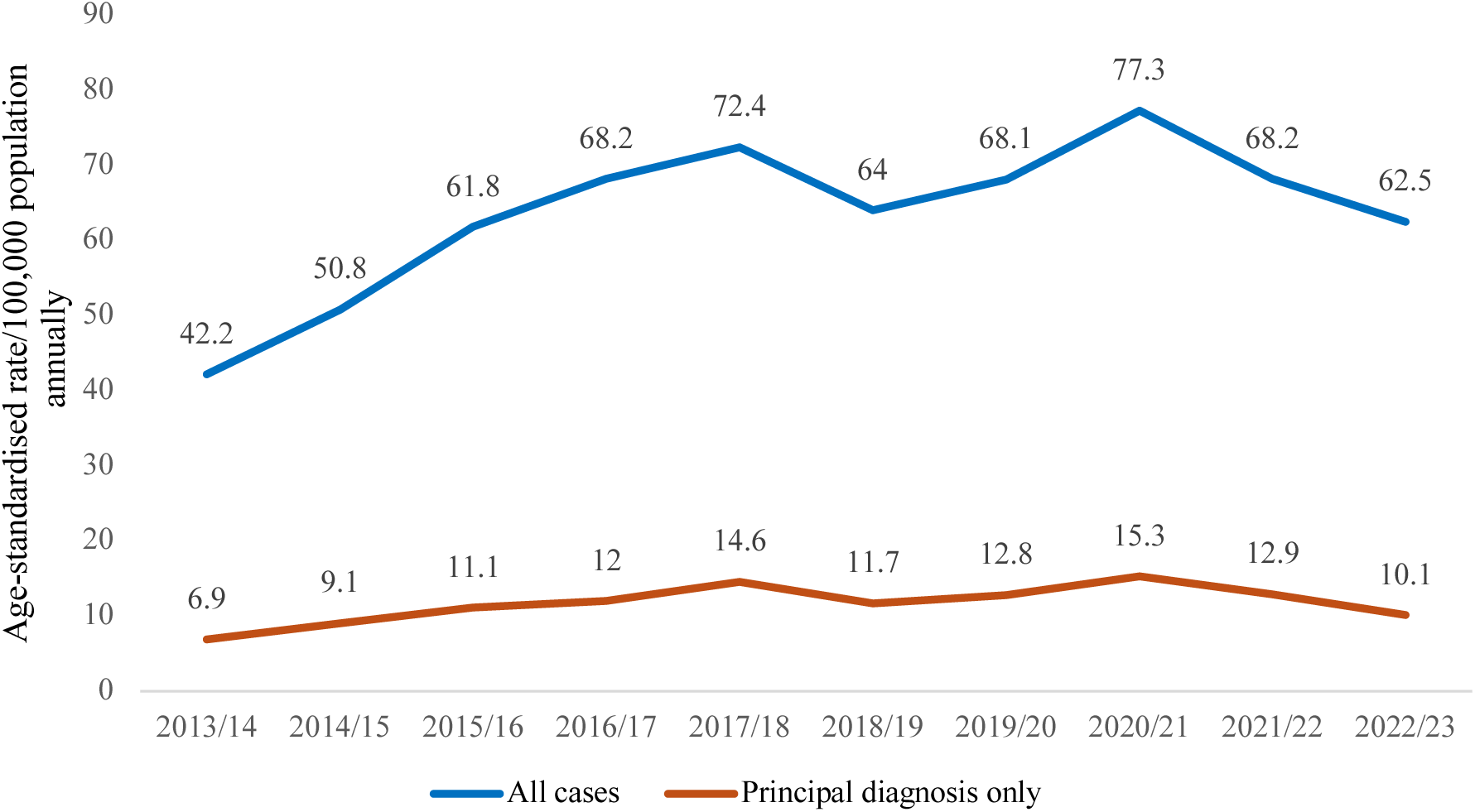
Cannabis-related hospital admission rate in Victoria, Australia 2013/14 to 2022/23: Age-standardised rates for all cases and principal diagnosis only cases

Most cases had a mental health and behavioural disorder diagnosis (F120-F129) (n=37,821; 95.6%), with an age-standardised rate per 100,000 population annually of 60.8. The greatest proportion of cases were related to harmful cannabis use (n=23,230; 58.7%), followed by withdrawal (n=7,029; 17.8%), psychotic disorder (n=5,785; 14.6%) and dependence (n=2,779; 7.0%). For cannabis-related cases overall, intoxication (n=1,839; 4.6%) and poisonings (n=1,859; 4.7%) each accounted for less than 5% of cases.

For admissions where cannabis-related harm was the principal diagnosis (n=7,263), mental and behavioural disorders were also the most common diagnosis (n=6,421; 88.4%). In these admissions, psychotic disorder was the main cannabis-related principal diagnosis (n=2,909; 40.1%), followed by harmful use (n=1,103; 15.2%). A similar proportion of cases were reported for withdrawal (n=840; 11.6%), poisonings (n=842; 11.6%) and intoxication (n=811; 11.2%). Dependence was the principal cannabis-related diagnosis in 8.5% of cases (n=615). Annual age adjusted rates per 100,000 population were calculated for all cases and principal diagnosis cases are presented in Figure 2 and frequencies in Supplementary Table S1.).

**Figure 2.**
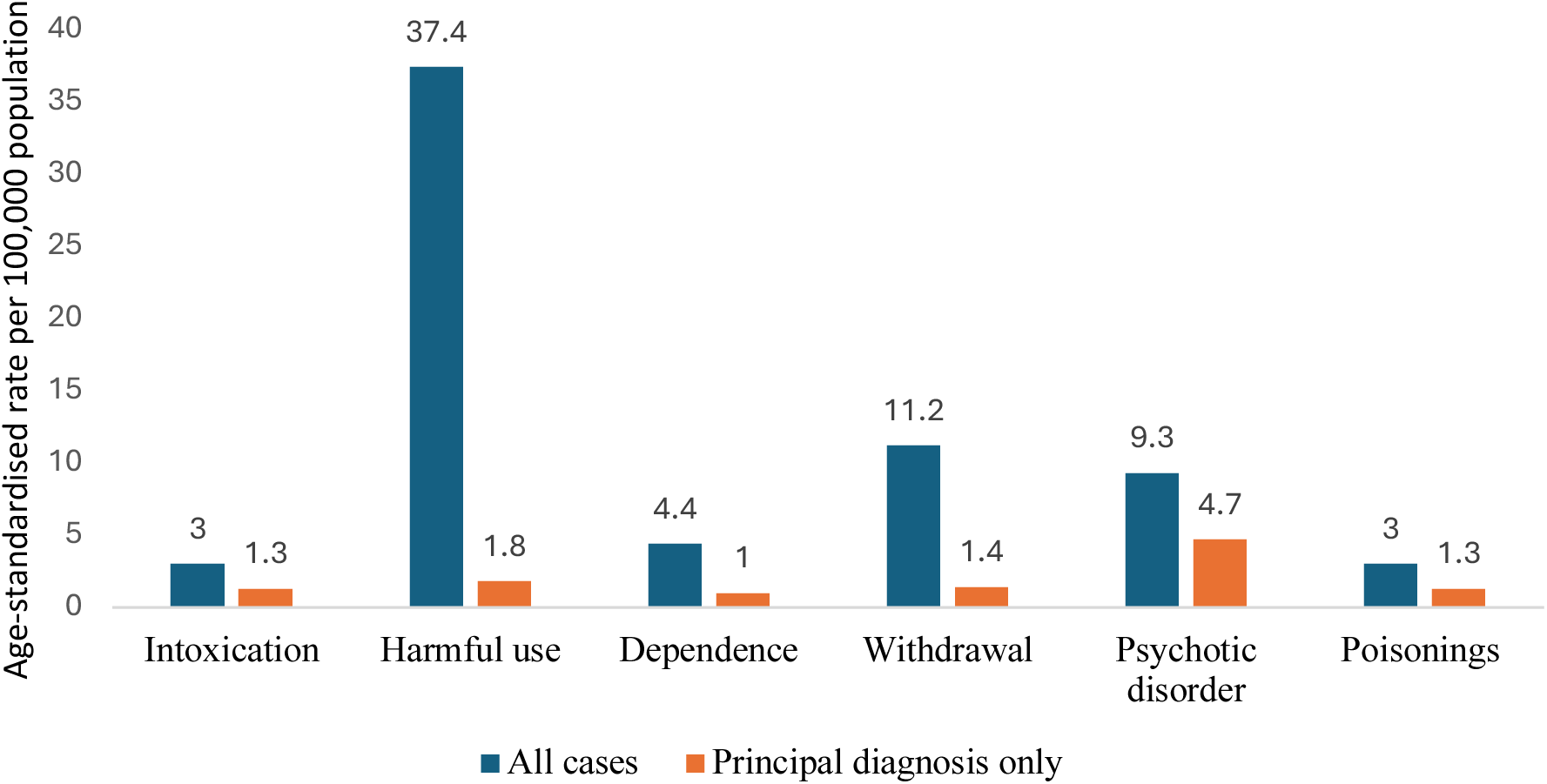
Cannabis-related hospital admissions in Victoria, Australia 2013/14 to 2022/23: Age-standardised rates per diagnosis code

### Admissions with a principal diagnosis that was not cannabis-related

To further differentiate between all cannabis-related cases and principal diagnosis only cases, admissions with a principal diagnosis that was not cannabis related were further examined. The top 12 most commonly occurring principal diagnoses (at 3-character level in the ICD-10- AM coding system) for cannabis-related hospital admissions where the principal diagnosis was not cannabis-related are listed in Table 1. These top 12 diagnoses accounted for over 62% of the cases.

**Table 1.** Admissions with a principal diagnosis that was not cannabis-related.

| ICD-10-AM code | Diagnosis | Frequency | Percent |
| --- | --- | --- | --- |
| F20 | Schizophrenia | 3999 | 12.38 |
| F15 | Mental and behavioural disorders due to use of other stimulants, including caffeine | 2683 | 8.31 |
| R11 | Nausea and vomiting | 2123 | 6.57 |
| F25 | Schizoaffective disorders | 2011 | 6.23 |
| F10 | Mental and behavioural disorders due to use of alcohol | 1889 | 5.85 |
| F31 | Bipolar affective disorder | 1809 | 5.60 |
| F60 | Specific personality disorders | 1488 | 4.61 |
| F32 | Depressive episode | 1234 | 3.82 |
| F43 | Reaction to severe stress, and adjustment disorders | 1121 | 3.47 |
| F29 | Unspecified nonorganic psychosis | 714 | 2.21 |
| R45 | Symptoms and signs involving emotional state | 620 | 1.92 |
| F41 | Other anxiety disorders | 493 | 1.53 |

Overall, of the cannabis-related hospital admissions where the principal diagnosis was not cannabis-related (n=32,302/39,565), the principal diagnosis was most often mental and behavioural disorders (ICD-10-AM “F” chapter, 61.5%), followed by general symptoms and signs (“R” chapter, 12.5%), injury, poisoning and certain other consequences of external causes (“S”/”T” chapters, 7.8%), pregnancy, childbirth and the puerperium (“O” chapter, 3.1%), diseases of the respiratory system (“J” chapter, 2.9%), diseases of the digestive system (“K” chapter, 2.4%), diseases of the circulatory system (“I” chapter, 2.0%) or other (7.7%).

### Sociodemographic characteristics

Males and females aged 15-24 years had the highest annual age-standardised rates per 100,000 population (145 and 110, respectively) and for those with cannabis-related harm as the principal diagnosis (36.9 and 20.5, respectively) (See Figure 3 for rates and Supplementary Table S2. for frequencies). Although case numbers and rates were higher in males, the adjusted net relative change in the 2022/23 rate compared with the 2013/14 rate was greater in females than in males (+68.7% vs. +39.4% for females vs. males, for all cannabis-related cases; +75.1% vs. +34.6% for females vs. males for principal diagnosis cases). The greatest adjusted net change in rate in 2022/23 compared with 2013/14 occurred in older adults aged 65+ years of both sexes (males +252.7% and females +411.3%); however, noting that the proportion of older adults was less than 1% of the overall sample (See Appendix Table A2).

**Figure 3.**
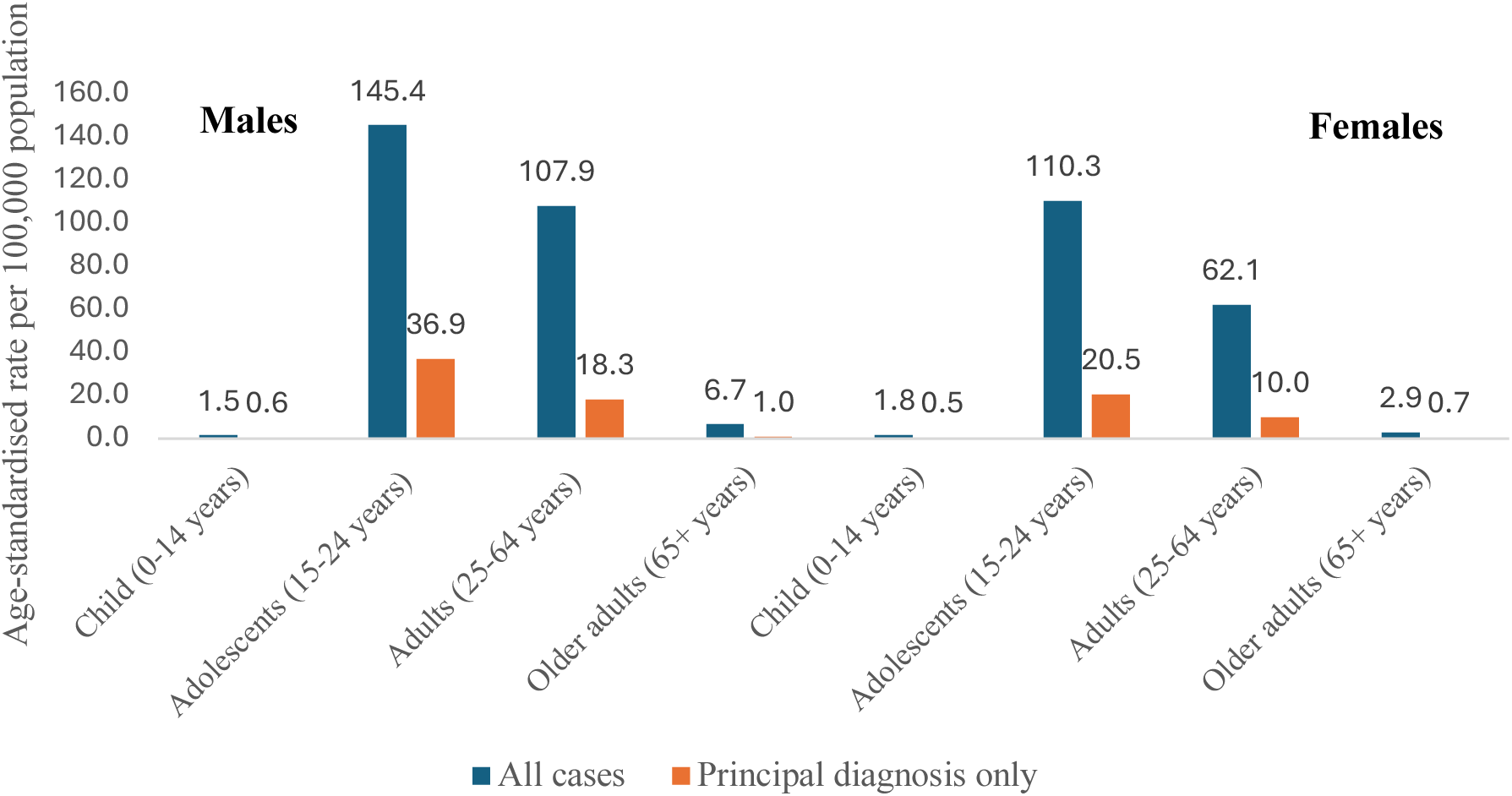
Cannabis-related hospital admission rates by age category and sex 2013/14 to 2022/23

### Intensive care unit (ICU) and overall hospital length of stay

There were n=1,592/39,565 (4.0%) cases with ICU stay of one hour or longer. ICU stay in admissions with a cannabis-related principal diagnosis occurred in 105/7,263 (1.5%) cases. The n=1,592 admissions overall with ICU stay had a total sum of 109,881 ICU hours. The n=105 admissions with a cannabis-related principal diagnosis and any ICU stay had a total sum of 5,411 ICU hours.

Overall, n=7,285 (18.4%) admissions were same-day admissions, n=5,552 (14.0%) were overnight admissions, and n=26,728 (67.6%) were multi-day admissions with a median of 7 days. Of the admissions with a cannabis-related principal diagnosis, n=1,926 (26.5%) were same-day admissions, n=1,126 (15.5%) were overnight admissions, and n=4,211 (57%) were multi-day admissions with a median of 7 days. It should be noted that these are incident admissions (i.e., these statistics do not include subsequent transfers or changes in care type within the same hospital).

### Discharge destination and deaths

Of the admissions for all cases, and those with a cannabis-related principal diagnosis, the majority (n=34,527/39,565; 87.3% and n=6,595/7,263; 90.8%, respectively) returned to private residence/accommodation. Over the ten-year period, there were n=53 deaths, and for cases where there was a cannabis-related principal diagnosis, there were <5 deaths (see Appendix A3).

## DISCUSSION

Comparing 2013/14 and 2022/23, there was a 49% increase in the rate of cannabis-related hospitalisations in Victoria, Australia. Notably, the time trend was nonlinear, with peaks at 2017/2018 and 2020/21 time points, with lower rates in the following 2 years. Less than 1 in 5 of all cannabis-related hospital admissions involved a cannabis-related principal diagnosis. Mental and behavioural disorder diagnoses were prominent in principal diagnosis cases, particularly psychotic disorder. Mental health comorbidities and polysubstance use were evident in cases with a principal diagnosis that was not cannabis related. Over two-thirds of cannabis-related principal diagnosis hospitalisations involved multi-day admissions. Hospitalisations were more common in adolescents and young people. While cases involving males are numerically greater, the largest percentage change over the 10 years was for females aged 15-24 years.

North American and European studies have examined cannabis-related hospitalisations in relation to policy changes or trends over time, with most studies reporting an increase in cannabis-related hospitalisations following policies with provisions for increased access (12–18). Australia offers a unique case study, with a decade of federally regulated medical access and no legal recreational market, yielding insights with global relevance. Consistent with worldwide data, cannabis remains the most consumed illegal drug in Australia. There are limited detailed studies that examine cannabis-related hospitalisation trends in Australia over the last decade (19).

Understanding the rate of cannabis-related hospitalisations in Australia is important from both a public health perspective and to determine the associated health system burdens. Principal diagnosis of cannabis-related hospitalisations is in the top five drug classes and precedes opioid-related hospitalisations (20). A key finding based on our study outcomes was that two-thirds of the cannabis-related admissions involved multi-day stays. This contrasts with national public hospital admissions, with nearly two-thirds being same-day separations (19). Although emergency department presentations are typically greater in number, hospital admissions are likely to reflect presentations that required additional treatment or monitoring (21). A 2015/16 study estimated inpatient hospital care costs at $123,511,008 and, more broadly across the healthcare system, at more than $700 million (22). While outside the scope of this study, future research could determine the healthcare system costs from 2016 onwards.

There are a range of possible reasons why cannabis harms may have increased over the study period outside of policy or product THC strength changes. During the COVID-19 pandemic, there were higher than forecasted national cannabis-related hospitalisation rates (23). In 2020, there were declines in emergency presentations and elective procedures (24, 25). Between 2021 and 2022, there was a substantially higher threshold for hospitalisations (i.e., acute, life-threatening cases) (26). The impacts of the extensive lockdown periods on mental health, income loss and social isolation could have influenced both substance use and healthcare system utilisation during this period. Young people and women were more severely impacted (27, 28). There are some parallels between these impacted populations and significant increases in cannabis-related hospitalisations over the decade in young females in our study findings. During this same period, domestic cultivation of cannabis meant that, unlike several other drugs, the supply chain was relatively uninterrupted by the COVID-19 pandemic, leading to shifts in substance use. Cannabinoid wastewater loads were also high during this period (29). These factors are consistent with the peak in cannabis-related hospitalisation age-standardised rate per 100,000 population annually that was observed in this study at the 2020/21 timepoint.

Sex- and age-based differences in prevalence of use during the same period provide additional insights. Findings in our study mirror national cannabis-related hospitalisation trends, with males accounting for the greatest number of cannabis related hospitalisations over the decade. Age-based comparisons between our state-based data and national data are limited by differences in reporting time periods, rate calculations, and age range categories (20). Similar demographic characteristics (i.e. males and/or adolescents and young adults) are evident in international cannabis-hospitalisation literature (18, 30, 31).

Increased hospitalisations among female adolescents and adults have narrowed the gap between male and female hospitalisations (4). The significant percentage increase in cannabis-related hospitalisations in young females observed in our study coincides with increased prevalence of past year cannabis use in females, rising from 7.6% to 9.8% over the study period (4). Given the potential harms of cannabis use in individuals less than 25 years of age and females of childbearing age (32), these trends provide insights into demographic sub-groups who may benefit from evidence-based education or public health initiatives.

### Limitations

As with all studies using administrative data, there are known limitations to acknowledge. Despite standardised ICD-10-AM coding, it is important to acknowledge that patterns of diagnosis and coding behaviour can change over time. Cannabis-related harms are likely to be underestimated in hospitalisation data, due to patient concerns about stigma or repercussions influencing use disclosure, and clinician screening and recording practices. It is unknown to what extent changing population attitudes and perceptions of cannabis influence their comfort in disclosure to healthcare providers in acute care settings. The rates presented are for admissions rather than individual people, and it is possible that the same person may have had multiple episodes of care. Multiple episodes, even if by the same person, will contribute to a higher population-based rate (33). Future research examining other measures of harm, such as ambulance attendances and emergency department presentations, may help generate a more comprehensive picture of cannabis related harm by leveraging the unique strengths and limitations of each data source. For example, hospital admission data may provide a more accurate insight into poisoning-related presentations and older populations than emergency department presentations (21). Longer term harms, such as cannabis use disorder, may also not be well captured (34).

Cannabis-related hospitalisation data does not include information on use motivation (i.e., medical and/or nonmedical), cannabinoid composition, dosage (i.e., frequency and quantity), dosage form, route of administration, or source (i.e., prescribed, diverted, or illicit). Synthetic cannabinoid or higher THC products that have distinct safety profiles cannot be differentiated. Polypharmacy was not examined in this study, including interactions between cannabinoids and other medications and substances patients were taking. While Victoria, Australia, has the most comprehensive capture of hospitalisation data, the findings of this study are not generalisable to other Australian states and territories. Despite these limitations, monitoring harms related to cannabis remains important from a public health perspective.

### Future directions

The prevalence of cannabis consumption in relation to the proportion of the Australian population experiencing harms is a critical consideration when interpreting these study outcomes. It is unknown to what extent the increased volume of higher THC medicinal cannabis prescription use translates into prescriptions dispensed or undesirable patient outcomes. Once additional years of hospitalisation data become available, this study could serve as a baseline from which to examine trends following higher THC inhaled product and edible prescription product applications. Additional insights could be gained by future studies that collect more detailed case information and analyse blood and product cannabinoid concentrations when an individual presents or is admitted for a cannabis-related harm. These findings point to rising acute mental health harms, particularly psychotic disorders, in the context of rising medical use and broader increases in cannabis-related harms. This flags the need for targeted research and clinical attention to this emerging public health concern, especially as cannabis use and use disorder represent modifiable risk factors for mental illness.

## CONCLUSION

Our findings demonstrate an overall 49% increase in cannabis-related hospital admissions over a decade, with a non-linear pattern with peaks in 2017/18 and 2020/21. Mental health and behavioural disorder presentations, such as psychotic disorder, are prominent in principal diagnosis cases. Influences on cannabis-related hospitalisation trends are likely to be multifactorial and include the influence of changes in cultural and social norms. Increases in cannabis-related hospitalisation trends in youth and young people underscore the need to implement evidence-based public health measures to reduce potential harms.

## Data Availability

All data produced in the present work are contained in the manuscript

**Table A1.** Age-standardised rate per 100,000 population annually for all cannabis-related cases and principal diagnosis only.

|  | All cases |  | Principal diagnosis only |  |
| --- | --- | --- | --- | --- |
|  | N (%) | Age-standardised rate per 100,000 pop annually | N (%) | Age-standardised rate per 100,000 pop annually |
| **Year** |  |  |  |  |
| 2013/14 | 2,420 (6.1%) | 42.2 | 394 (5.4%) | 6.9 |
| 2014/15 | 2,969 (7.5%) | 50.8 | 537 (7.4%) | 9.1 |
| 2015/16 | 3,688 (9.3%) | 61.8 | 662 (9.1%) | 11.1 |
| 2016/17 | 4,167 (10.5%) | 68.2 | 743 (10.2%) | 12.0 |
| 2017/18 | 4,533 (11.5%) | 72.4 | 917 (12.6%) | 14.6 |
| 2018/19 | 4,085 (10.3%) | 64.0 | 746 (10.3%) | 11.7 |
| 2019/20 | 4,410 (11.1%) | 68.1 | 831 (11.4%) | 12.8 |
| 2020/21 | 4,887 (12.4%) | 77.3 | 964 (13.3%) | 15.3 |
| 2021/22 | 4,317 (10.9%) | 68.2 | 811 (11.2%) | 12.9 |
| 2022/23 | 4,089 (10.3%) | 62.5 | 658 (9.1%) | 10.1 |
| Total 2013/14 to 2022/23 | 39,565 (100.0%) | 63.6 | 7,263 (100.0%) | 11.7 |
| **Males** |  |  |  |  |
| 2013/14 | 1,507 (6.2%) | 53.3 | 253 (5.4%) | 9.0 |
| 2014/15 | 1,944 (8.0%) | 67.3 | 370 (7.9%) | 12.6 |
| 2015/16 | 2,331 (9.5%) | 79.2 | 437 (9.3%) | 14.8 |
| 2016/17 | 2,665 (10.9%) | 88.1 | 509 (10.9%) | 16.6 |
| 2017/18 | 2,872 (11.8%) | 93.1 | 626 (13.3%) | 20.1 |
| 2018/19 | 2,591 (10.6%) | 81.7 | 469 (10.0%) | 14.8 |
| 2019/20 | 2,759 (11.3%) | 85.9 | 548 (11.7%) | 17.0 |
| 2020/21 | 2,883 (11.8%) | 91.8 | 612 (13.0%) | 19.6 |
| 2021/22 | 2,498 (10.2%) | 79.6 | 484 (10.3%) | 15.6 |
| 2022/23 | 2,373 (9.7%) | 73.1 | 383 (8.2%) | 11.8 |
| Total for males 2013/14 to 2022/23 | 24,423 (100%) | 79.3 | 4,691 (100%) | 15.2 |
| **Females** |  |  |  |  |
| 2013/14 | 913 (6.0%) | 31.4 | 141 (5.5%) | 4.8 |
| 2014/15 | 1,025 (6.8%) | 34.7 | 167 (6.5%) | 5.7 |
| 2015/16 | 1,357 (9.0%) | 45.0 | 225 (8.7%) | 7.6 |
| 2016/17 | 1,502 (9.9%) | 48.9 | 234 (9.1%) | 7.6 |
| 2017/18 | 1,661 (11.0%) | 52.4 | 291 (11.3%) | 9.3 |
| 2018/19 | 1,494 (9.9%) | 46.8 | 277 (10.8%) | 8.8 |
| 2019/20 | 1,651 (10.9%) | 50.9 | 283 (11.0%) | 8.6 |
| 2020/21 | 2,004 (13.2%) | 63.3 | 352 (13.7%) | 11.1 |
| 2021/22 | 1,819 (12.0%) | 57.2 | 327 (12.7%) | 10.4 |
| 2022/23 | 1,716 (11.3%) | 52.3 | 275 (10.7%) | 8.4 |
| Total for females 2013/14 to 2022/23 | 15,142 (100%) | 48.3 | 2,572 (100%) | 8.2 |

**Table A2.** Adjusted net percentage change in cannabis-related admission rates from baseline (2013/14) to the final study year (2022/23) for all cases and for cases with a cannabis-related principal diagnosis only.

|  | All cases |  |  |  | Principal diagnosis only |  |  |  |
| --- | --- | --- | --- | --- | --- | --- | --- | --- |
|  | Adjusted total % change (2013/14 to 2022/23)* | Lower CL | Upper CL | p-value | Adjusted total % change (2013/14 to 2022/23)* | Lower CL | Upper CL | p-value |
| Overall | 50.35 | 35.58 | 66.72 | <.0001 | 49.03 | 26.47 | 75.61 | <.0001 |
| **Age group x sex** |  |  |  |  |  |  |  |  |
| *Male* | 39.44 | 24.08 | 56.69 | <.0001 | 34.6 | 10.48 | 63.99 | 0.0032 |
| Child (0-14 years) | 155.55 | -19.65 | 712.73 | 0.112 | 161.25 | -40.69 | 1050.9 | 0.2043 |
| Adolescents (15-24 years) | 17.08 | -0.31 | 37.5 | 0.0545 | 21.44 | -12.6 | 68.75 | 0.2471 |
| Adults (25-64 years) | 44.64 | 22.67 | 70.55 | <.0001 | 36.92 | 3.2 | 81.65 | 0.0294 |
| Older adults (65+ years) | 252.71 | 59.93 | 677.85 | 0.0018 | † |  |  |  |
| *Females* | 68.68 | 44.72 | 96.61 | <.0001 | 75.14 | 34.63 | 127.84 | <.0001 |
| Child (0-14 years) | -25.45 | -69.54 | 82.47 | 0.5202 | † |  |  |  |
| Adolescents (15-24 years) | 88.63 | 48.3 | 139.94 | <.0001 | 122.1 | 35.95 | 262.84 | 0.0014 |
| Adults (25-64 years) | 59.9 | 26.25 | 102.54 | <.0001 | 57.98 | 6.98 | 133.28 | 0.0215 |
| Older adults (65+ years) | 411.34 | 61.86 | 1515.38 | 0.0054 | 214.31 | -63.55 | 2610.3 | 0.2975 |

\*Estimated from generalised linear models (Poisson distribution with log-population offset) with financial year entered as a categorical variable. Percentage represents the net relative change in the 2022/23 rate compared to the 2013/14 baseline, adjusted for age group and sex.
†Estimates could not be calculated for this subgroup due to model non-convergence resulting from sparse data.

**Table A3.** Discharge destination for cannabis-related cases.

| **Separation mode** | **Frequency** | **Percent** |
| --- | --- | --- |
| Separation and transfer to mental health residential facility | 785 | 1.98 |
| Separation and transfer to Transition Care bed-based program | 29 | 0.07 |
| Death | 53 | 0.13 |
| Separation to private residence/accommodation | 34527 | 87.27 |
| Statistical Separation (change in Care Type within this hospital) | 866 | 2.19 |
| Separation and transfer to other acute hospital/extended care/rehabilitation/geriatric centre | 1416 | 3.58 |
| Left against medical advice | 1863 | 4.71 |
| Other | 26 | 0.07 |

## Supplementary Files

**Table S1.** Cannabis-related hospital admissions in Victoria, Australia 2013/14 to 2022/23: frequencies per diagnosis code.

| **Diagnosis code** | **All cases** | **Principal diagnosis only** |
| --- | --- | --- |
| Intoxication | 1,839 | 811 |
| Harmful use | 23,230 | 1,103 |
| Dependence | 2,779 | 615 |
| Withdrawal | 7,029 | 840 |
| Psychotic disorder | 5,785 | 2,909 |
| Poisonings | 1,859 | 842 |

**Table S2.**
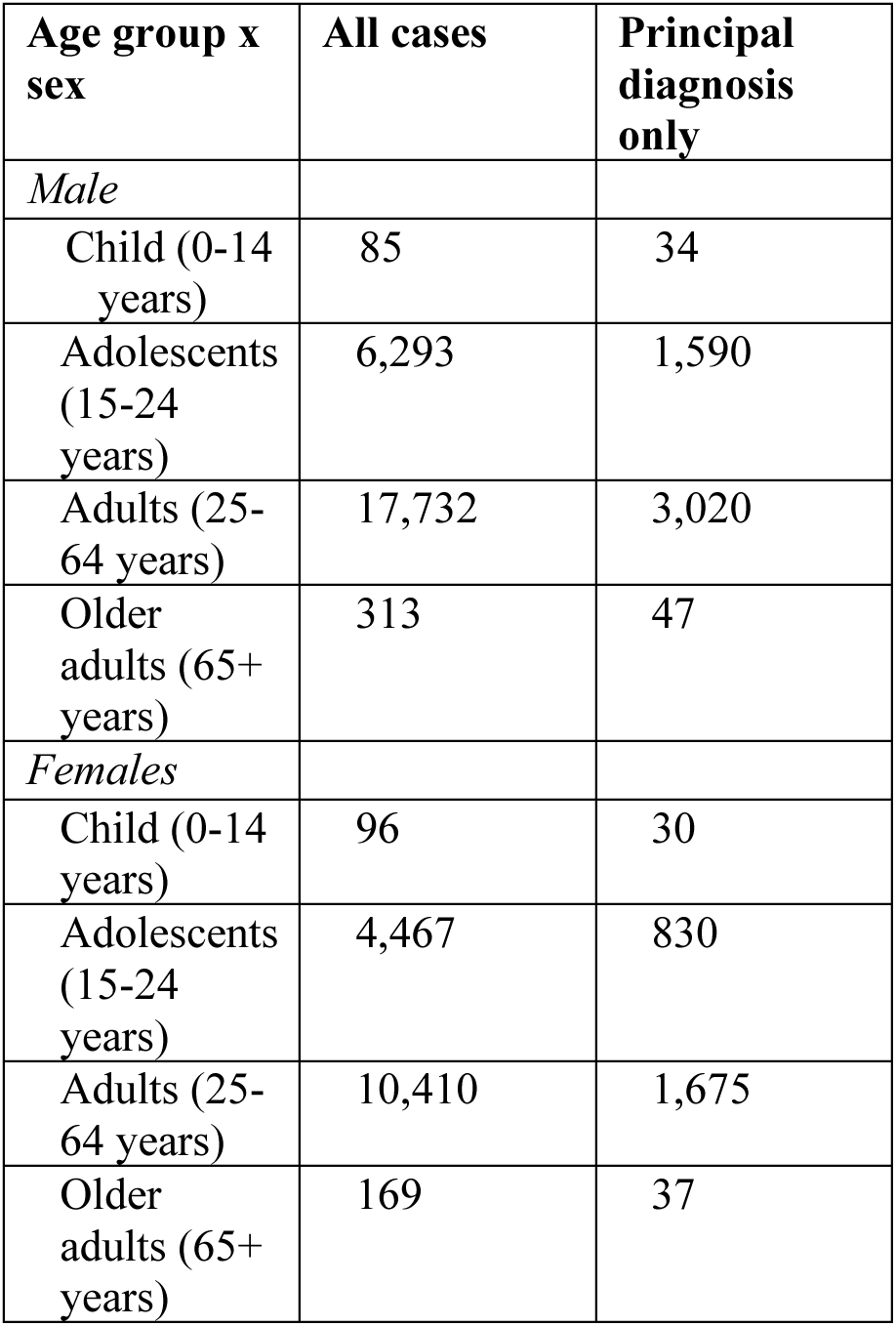
Cannabis-related hospital admission frequencies by age category and sex 2013/14 to 2022/23.

| **Age group x sex** | **All cases** | **Principal diagnosis only** |
| --- | --- | --- |
| *Male* |  |  |
| Child (0-14 years) | 85 | 34 |
| Adolescents (15-24 years) | 6,293 | 1,590 |
| Adults (25-64 years) | 17,732 | 3,020 |
| Older adults (65+ years) | 313 | 47 |
| *Females* |  |  |
| Child (0-14 years) | 96 | 30 |
| Adolescents (15-24 years) | 4,467 | 830 |
| Adults (25-64 years) | 10,410 | 1,675 |
| Older adults (65+ years) | 169 | 37 |

**Figure S1.**
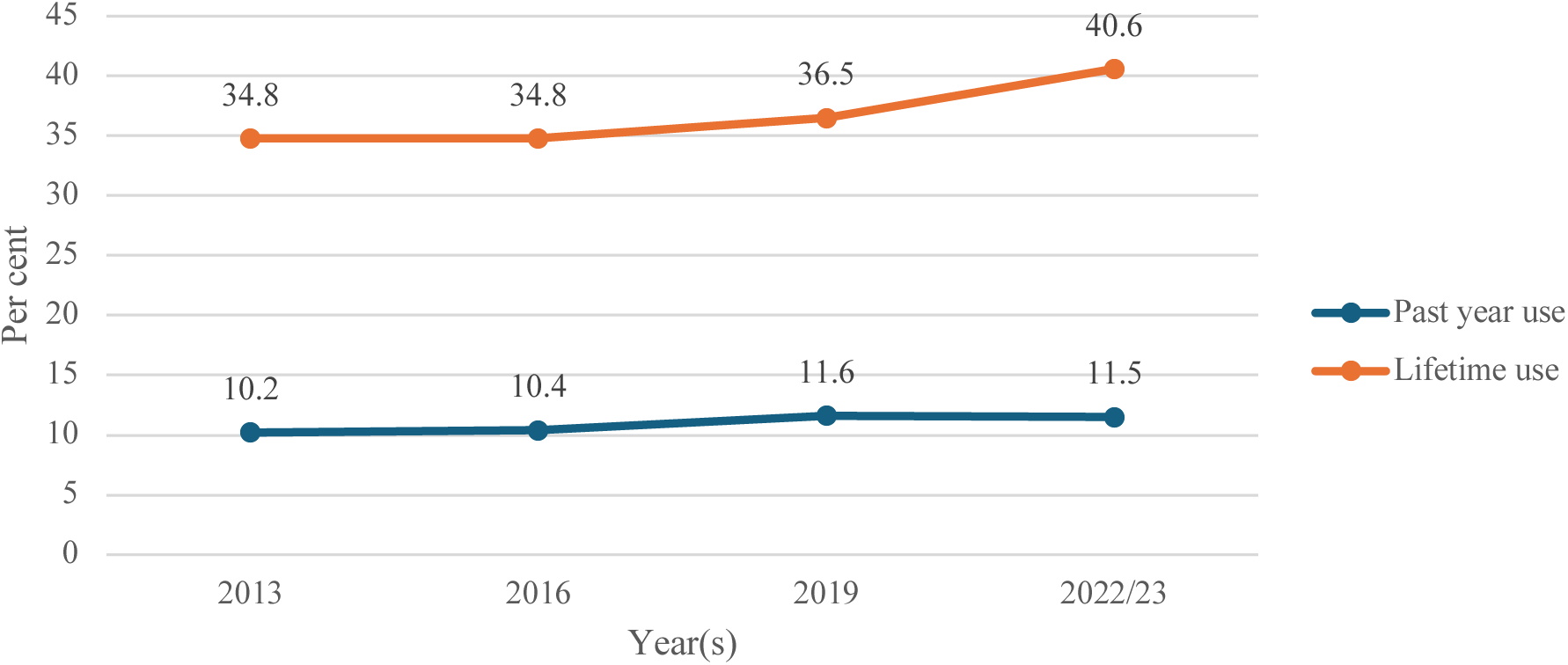
Cannabis use trends in the Australian population between 2013 and 2023 (data source AIHW 2026)

**Figure S2.**
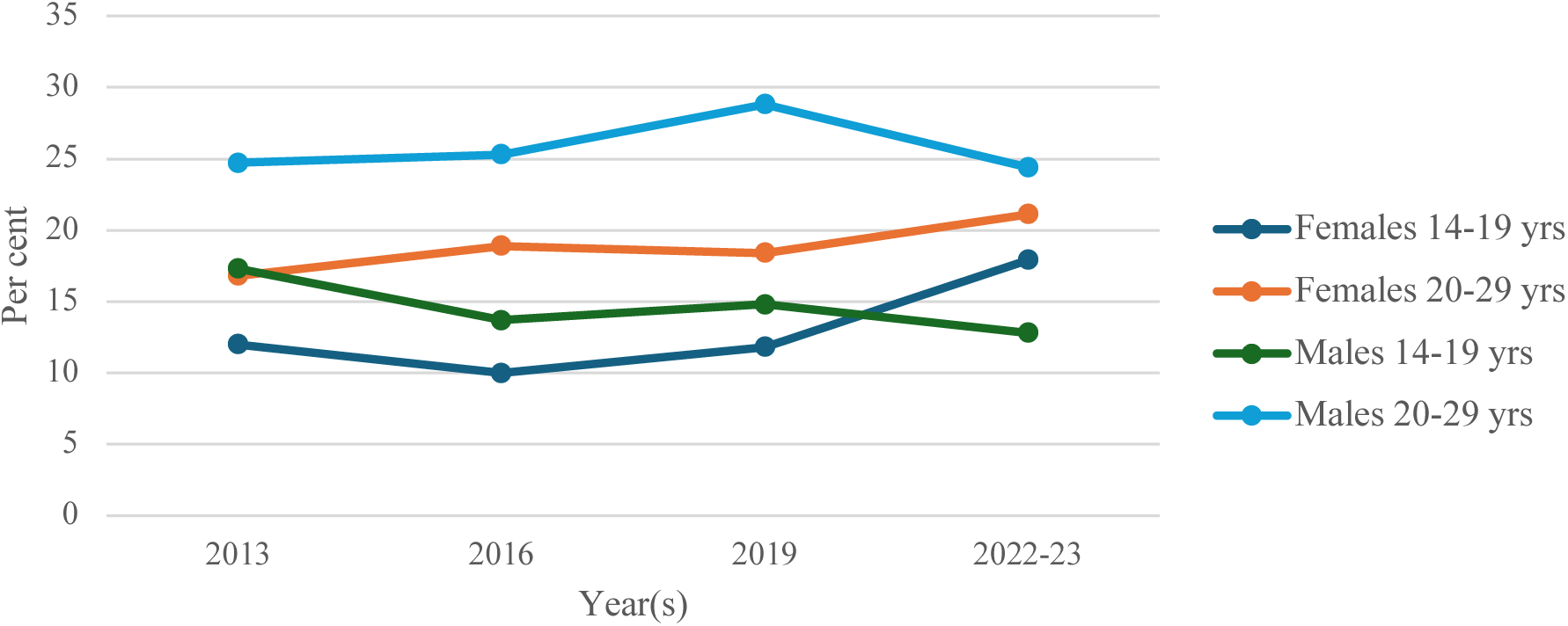
Past year cannabis use by adolescents and younger people, by age category and gender, 2013 to 2022-23 (data source AIHW 2026)

